# Biometric Contrastive Learning for Data-Efficient Deep Learning from Electrocardiographic Images

**DOI:** 10.1101/2023.09.13.23295494

**Authors:** Veer Sangha, Akshay Khunte, Gregory Holste, Bobak J Mortazavi, Zhangyang Wang, Evangelos K Oikonomou, Rohan Khera

**Author notes:** **Address for correspondence:** Rohan Khera, MD, MS 195 Church St, 6th Floor, New Haven, CT 06510; @rohan_khera.

## Abstract

**Objective:** Artificial intelligence (AI) detects heart disease from images of electrocardiograms (ECGs), however traditional supervised learning is limited by the need for large amounts of labeled data. We report the development of Biometric Contrastive Learning (BCL), a self-supervised pretraining approach for label-efficient deep learning on ECG images.

**Materials and Methods:** Using pairs of ECGs from 78,288 individuals from Yale (2000-2015), we trained a convolutional neural network to identify temporally-separated ECG pairs that varied in layouts from the same patient. We fine-tuned BCL-pretrained models to detect atrial fibrillation (AF), gender, and LVEF<40%, using ECGs from 2015-2021. We externally tested the models in cohorts from Germany and the US. We compared BCL with random initialization and general-purpose self-supervised contrastive learning for images (simCLR).

**Results:** While with 100% labeled training data, BCL performed similarly to other approaches for detecting AF/Gender/LVEF<40% with AUROC of 0.98/0.90/0.90 in the held-out test sets, it consistently outperformed other methods with smaller proportions of labeled data, reaching equivalent performance at 50% of data. With 0.1% data, BCL achieved AUROC of 0.88/0.79/0.75, compared with 0.51/0.52/0.60 (random) and 0.61/0.53/0.49 (simCLR). In external validation, BCL outperformed other methods even at 100% labeled training data, with AUROC of 0.88/0.88 for Gender and LVEF<40% compared with 0.83/0.83 (random) and 0.84/0.83 (simCLR).

**Discussion and Conclusion:** A pretraining strategy that leverages biometric signatures of different ECGs from the same patient enhances the efficiency of developing AI models for ECG images. This represents a major advance in detecting disorders from ECG images with limited labeled data.

## BACKGROUND AND SIGNIFICANCE

Electrocardiography is a ubiquitous tool for the diagnosis of cardiovascular diseases. Deep learning has been successfully applied to automate both the detection of disorders that are commonly discernable by physicians from electrocardiograms (ECGs),^1,2^ as well as those that traditionally require more specialized imaging modalities such as echocardiography or cardiac magnetic resonance imaging scans^3–5^.

While most existing artificial intelligence (AI) tools to automatically analyze ECGs rely on raw signal data, most end users do not have access to ECG signals. We have developed an approach that can interpretably diagnose conduction and rhythm disorders,^6^ as well as structural disorders^7^ from any layout of real-world 12-lead ECG images. Our image-based AI-ECG approach is applicable to various clinical settings, different hospitals, and data storage formats, representing an easily accessible and scalable approach to detect underdiagnosed cardiovascular disorders in at risk populations.

Despite the advantages of an image-based approach, like any AI approach, regardless of the modality, algorithmic training and development require large, labeled datasets. However, many clinical disorders have low prevalence with few examples in any individual dataset to develop algorithms designed for those conditions. This low prevalence of clinical labels is a key challenge for the development and generalizability of supervised learning approaches for ECG image models. For this reason, we developed a self-supervised learning approach to learn representations of ECG images that can serve as initializations for downstream finetuning on small, labeled datasets.

Self-supervised learning is designed to reduce the reliance on labeled data to develop models. The approach leverages unlabeled data to pretrain models before downstream fine-tuning on small, labeled datasets. It has been used in natural^8^ and medical image^9,10^ tasks, and has recently been applied to ECG signals.^11–13^

## OBJECTIVES

While self-supervised or contrastive learning has been done on real-world images of objects, 12-lead ECG images do not have an existing strategy to learn deeper features from unlabeled ECG images. Moreover, applications of self-supervised contrastive learning that have used raw ECG voltage data have not been designed to detect deep or hidden features of structural heart disease.

We have developed a few-shot, deep learning model development strategy – biometric contrastive learning (BCL) – in which our models are first trained to detect homologies of ECG features belonging to the same person, allowing for enhanced learning and detection of structural and functional abnormalities of the heart from any ECG image. Therefore, by developing a model that is designed to learn that two distinct ECGs belong to the same person, we develop an approach that builds a model that already recognizes key hidden feature that make two ECGs of a single person similar. This early training process can then be fine-tuned on a small number of labeled examples of any disease of interest, and the model is able to adapt to detection of the new disorder.

## MATERIALS AND METHODS

The study was reviewed by the Yale Institutional Review Board, which approved the study protocol and waived the need for informed consent as the study represents secondary analysis of existing data. The data cannot be shared publicly.

### Data Sources for Model Development

12-lead ECG signal waveform data from the Yale New Haven Hospital (YNHH) collected between 2000 and 2021 was used for the development and validation of BCL. These ECGs were recorded as standard 12-lead recordings sampled at a frequency of 500 Hz for 10 seconds. They were recorded on multiple different machines, and a majority were collected using Philips PageWriter machines and GE MAC machines. ECGs from 2000-2015 were used to develop the pretraining model. No clinical or other labels were used for these ECGs.

ECGs collected between 2015 and 2020 were used to train supervised models on three clinical tasks. ECGs from separate patients collected in 2021 were used to evaluate the effectiveness of BCL and the other baseline methods.

### Data Preprocessing

ECG images were generated in the same manner as previous studies.^6,7^ Briefly, ECGs with 10 seconds of continuous recordings across all 12 leads were preprocessed with a one-second median filter, subtracted from the original waveform to remove baseline drift. ECG signals were transformed into ECG images using the Python library ecg-plot. All images were converted to greyscale, followed by down-sampling to 300×300 pixels regardless of their original resolution using Python Image Library (PIL v9.2.0).

We created datasets with different plotting schemes for each signal waveform recording to ensure models were adaptable to real-world images, which vary in formats and layouts of leads. Four formats of images were generated for use in both BCL model pretraining and classification fine-tuning tasks: 1) standard printed ECG format in the United States, with four 2.5-second columns and a lead I rhythm strip, 2) a two-rhythm strip format, added lead II as an additional rhythm strip to the standard format, 3) an alternate format which consisted of two columns, each with 5 seconds of recording, and 4) a shuffled format, which had precordial leads in the first two columns and limb leads in the third and fourth. All images were rotated a random amount between -10 and 10 degrees before being input into the model to mimic variations seen in uploaded ECGs and to aid in prevention of overfitting.

### Model Training Overview

BCL uses a convolutional neural network (CNN) backbone to build representations of ECGs specific to individuals. During pretraining, the model learned the elements of an ECG image that are consistent for a person. The model was rewarded for identifying ECG images from the same individual as similar and penalized for identifying ECG images from different individuals as similar.

### Positive and Negative Views

To develop the BCL model, we used ECG images in the four formats described above. Any two ECGs from the same person, in any two formats of image, were treated as a positive pair. Any pair of images from different individuals was treated as a negative pair.

### Alternative Approaches

In addition to BCL, we tested two traditional approaches for model pretraining. The first of these was random weight initialization. The second was the classic SimCLR^8^ contrastive pretraining method that has been popular in image processing, such as those to identify objects in real-world photos. The latter approach, developed by Google, uses cropped and flipped parts of an image to identify the parts derived from the same image.

### BCL Model Pretraining

We used the EfficientNet-B3 CNN architecture for our encoder, following the demonstrated effectiveness of the model on other ECG image classification tasks.^6,7^ This model architecture requires images to be sampled at 300 x 300 square pixels includes 384 layers and has over 10 million trainable parameters (**Figure S1**). ECG images were converted to greyscale before being input into the model. Of note, the method is not restricted to this CNN architecture.

Our goal during pretraining was to minimize the contrastive loss function, which depended on the cosine similarity of embeddings of positive pairs in each batch compared to embeddings of negative pairs in the same batch. We used batch sizes of 16, with each ECG in the batch having one positive pair and 14 negative pairs, and accumulated gradients over 16 batches before updating the model.

Our encoder had an output dimension of 1,536. We used a 2-layer multilayer perceptron (MLP) in pretraining, which projected the 1,536-dimensional output of the encoder into a 128-dimensional space. We used an Adam optimizer with a learning rate of 1×10^-5^ for pretraining and trained the model for 10 epochs.

### Dataset for BCL and SimCLR Pretraining

We pretrained our model using ECGs acquired between 2000 and 2015 without labels from the Yale New Haven Health System. We excluded patients with ECGs after 2015 to ensure there was no data leakage across pretraining and model development. For each patient, the pair of ECGs with the smallest time difference between 5 and 1000 days apart was chosen, and the other ECGs were not used. For each pair of ECGs, 6 unique pairs were created, corresponding to potential combinations where ECGs were in two different out of the 4 total image formats. During pretraining, we ensured that per batch there were no more than one pair of ECGs from any given patient.

### Downstream Classification Task Training

We performed downstream fine tuning and evaluation of our pretrained encoder on three clinical tasks: Atrial fibrillation (AF), gender, and left ventricular systolic dysfunction (LVSD) classification. They were chosen as they have different biological bases, and each has large prior work demonstrating their detection using ECGs.

AF is a rhythm disorder that is characterized by missing P waves and irregular heartbeats. AF is diagnosable by clinicians from ECGs upon manual inspection, and our previous work has demonstrated its diagnosis by AI algorithms for 12 lead signal and image data^1,2,6^. Cardiologist-confirmed diagnosis statements accompanying all ECGs in the development cohort were searched for strings referencing atrial fibrillation and its abbreviations to identify ECGs for the task.

In addition to AF, we chose two clinical labels on ECGs that cannot be inferred by even experts on ECGs. First, is the gender of the patient. Gender is a hidden label that is not discernable on ECGs, but previous work has suggested that it can be detected on ECGs using AI methods^1,6^. The second is LVSD, which is defined as LVEF < 40% is associated with over 8-fold increased risk of subsequent heart failure and 2-fold risk of premature death^14^. It is also a hidden label, that has traditionally been diagnosed using echocardiography. Recent work has shown that LVSD can be diagnosed using 12-lead ECGs^3,7,15^. ECGs in the development cohort within 15 days of a TTE were used for this task, and LVEF values from cardiologist’s read on the nearest TTE to each ECG were used.

We used the pretrained encoders described above and two randomly initialized fully connected layers to predict the labels of interest. We trained our classification models on progressively larger samples of training data, as 0.1%, 0.5%, 1%, 5%, 20%, 50%, and 100% labeled fractions of training datasets to test the effectiveness of various initialization methods. We trained with a learning rate of 1 x 10^-4^ for 40 epochs. We used an Adam optimizer, gradient clipping, and a minibatch size of 64 throughout training, and we stopped training when validation loss did not improve in 3 consecutive epochs.

### External Validation

We pursued external validation to assess the generalizability of these pretraining techniques to external data sources. In addition to the held-out test set, AF and gender models were validated in the Germany-Based dataset PTB-XL, whose data have previously been described^16^. The dataset has 21,837 recordings from 18,885 patients, which were collected between 1989 and 1996. LVSD models were validated in a deidentified dataset of inpatient admissions at Lake Regional Hospital (LRH) in Osage Beach, Missouri, which has also been previously described^7^. Briefly, the dataset contains 100 ECGs from unique patients, with 43 from patients with LVEF<40% as measured by a TTE within 15 days of the ECG.

### Statistical Analysis

Categorical variables were presented as frequency and percentages, and continuous variables as means and standard deviations or median and interquartile range, as appropriate. Model performance was evaluated in the held-out test set and external ECG image datasets. We used area under the receiver operator characteristic (AUROC) and the area under the precision-recall curve (AUPRC) to measure model discrimination. Analytic packages used in model development and statistical analysis are reported in **Table S1**. All model development and statistical analyses were performed using Python 3.9.5.

## RESULTS

### Study Population

ECGs from the Yale New Haven Health system were split into pretraining, development, and held-out test sets. Briefly, our pretraining dataset contained ECGs from 2000-2015, our development datasets for our AF, gender, and LVSD models used ECGs between 2015-2020, and our held-out test set used ECGs from January – June 2021. We constructed two development datasets, one that sampled all 382,830 ECGs that had a corresponding LVEF value from their nearest TTE within 15 days of recording to develop LVSD models. We then randomly sampled the same number, 382,830, from the total 1,869,582 ECGs recorded during this period to construct a development cohort for our AF and gender models. Similarly, we constructed equal sized temporally distinct held-out test sets from ECGs in 2021, each containing 8,708 ECGs, limited to one ECG per patient to ensure independence of observations in the assessment of performance metrics. **Table 1** describes the patient characteristics in the pretraining, development, and test cohorts. ECGs in the development cohort for all 3 classification tasks were split into training and validation datasets at the patient level (90% 10%), stratified by whether a patient had the condition of interest.

**Table 1.**
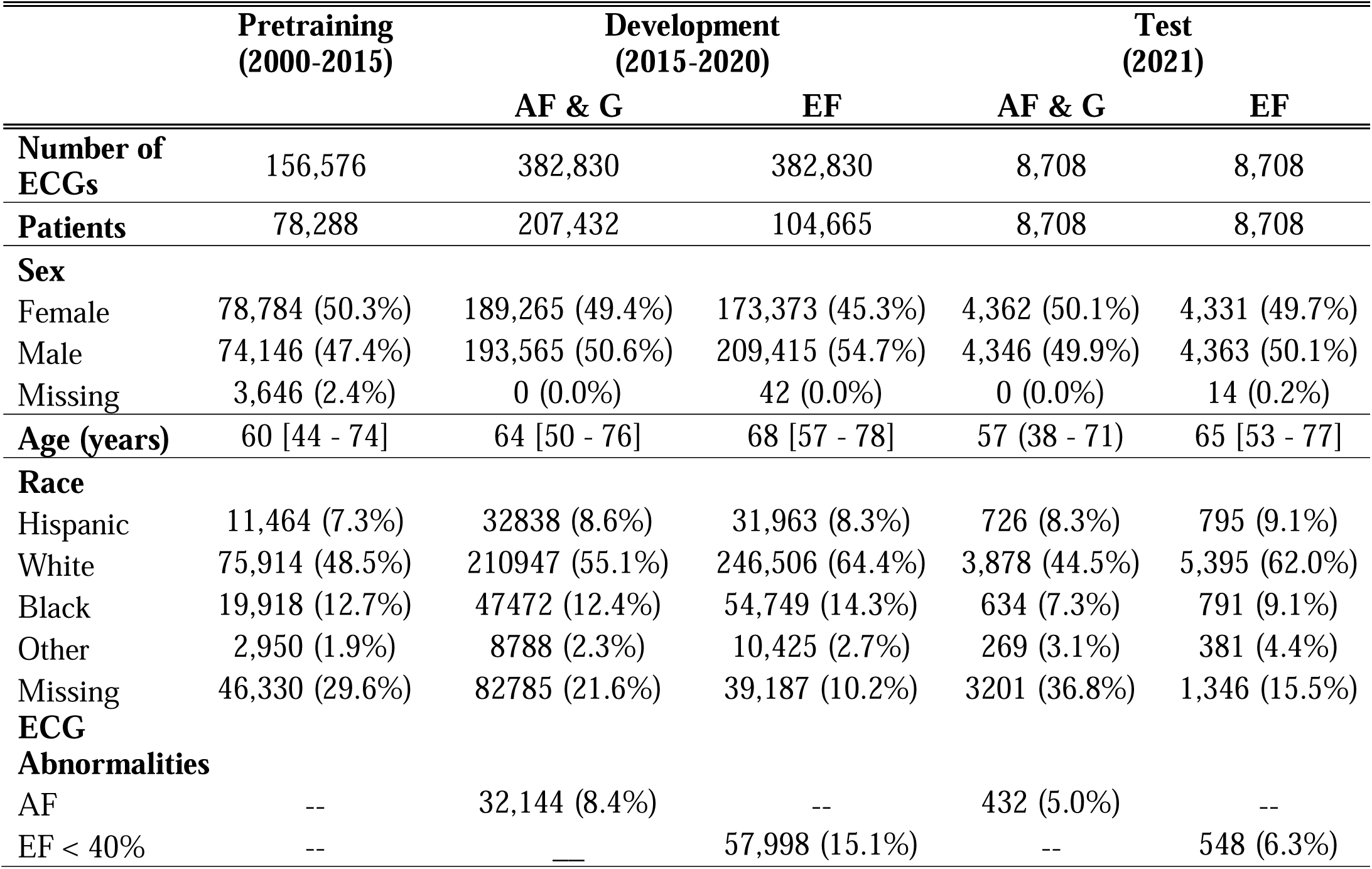
Baseline characteristics of study population. Data presented as median [IQR] for age and number (percent) for other variables. Abbreviations: ECGs, electrocardiograms; EF, ejection fraction; A-Fib, atrial fibrillation.

### Performance in the Held-Out Test Sets

When trained on 100% of the data available, the three strategies (BCL, random, simCLR) were comparable for the 3 tasks of identifying gender, atrial fibrillation, and LVSD, on both AUROC and AUPRC, the performance metrics we used to evaluate models. However, across the three tasks, BCL demonstrated equivalent performance for both AUROC (**Figure 2**) and AUPRC (**Figure 3**) at 50% of the data available as it did with 100% of the data, even though the other models initialized with random weights and using a SimCLR approach suffered drop-offs in performance.

**Figure 1.**
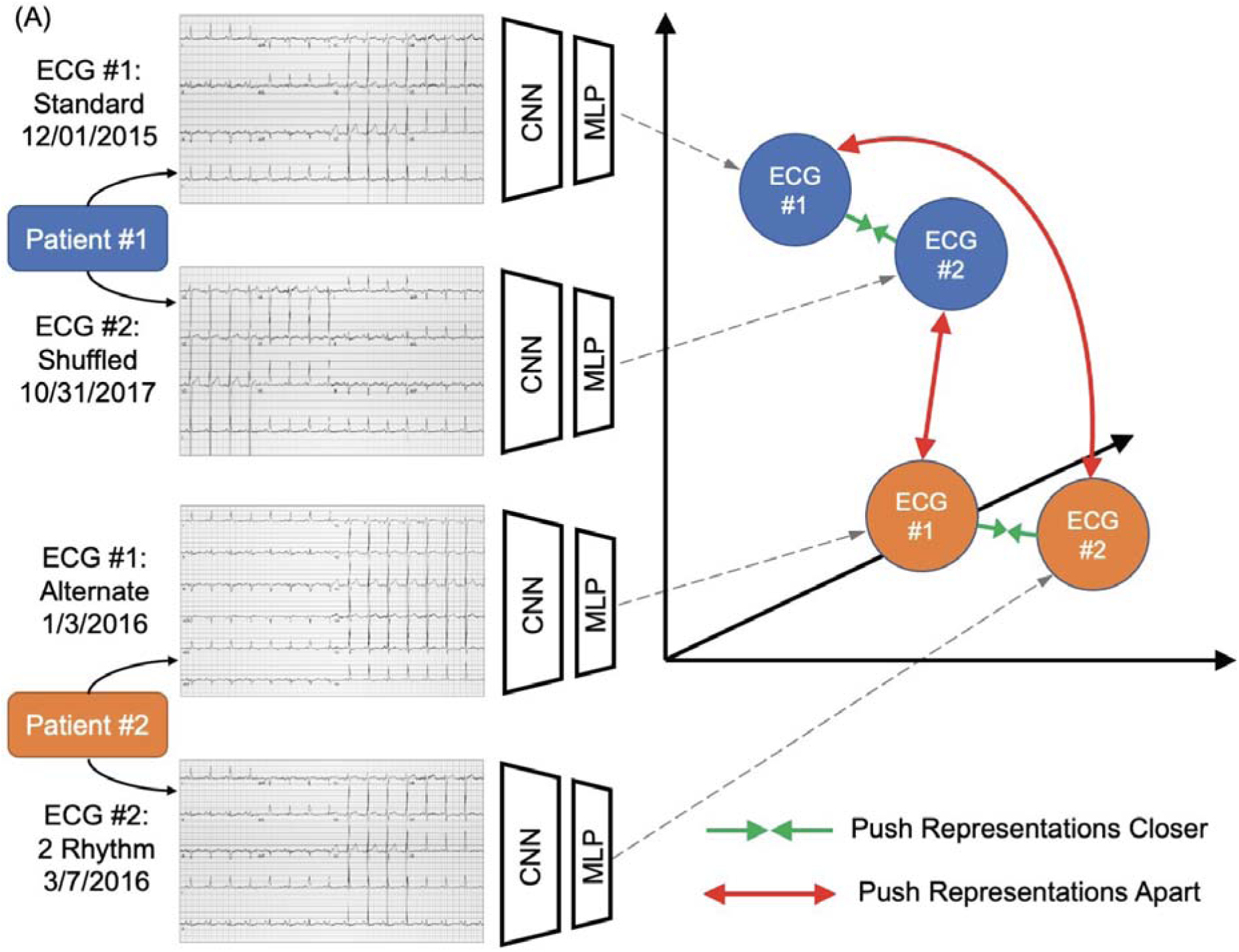
Overview of Biometric Contrastive Learning (BCL).

**Figure 2.**
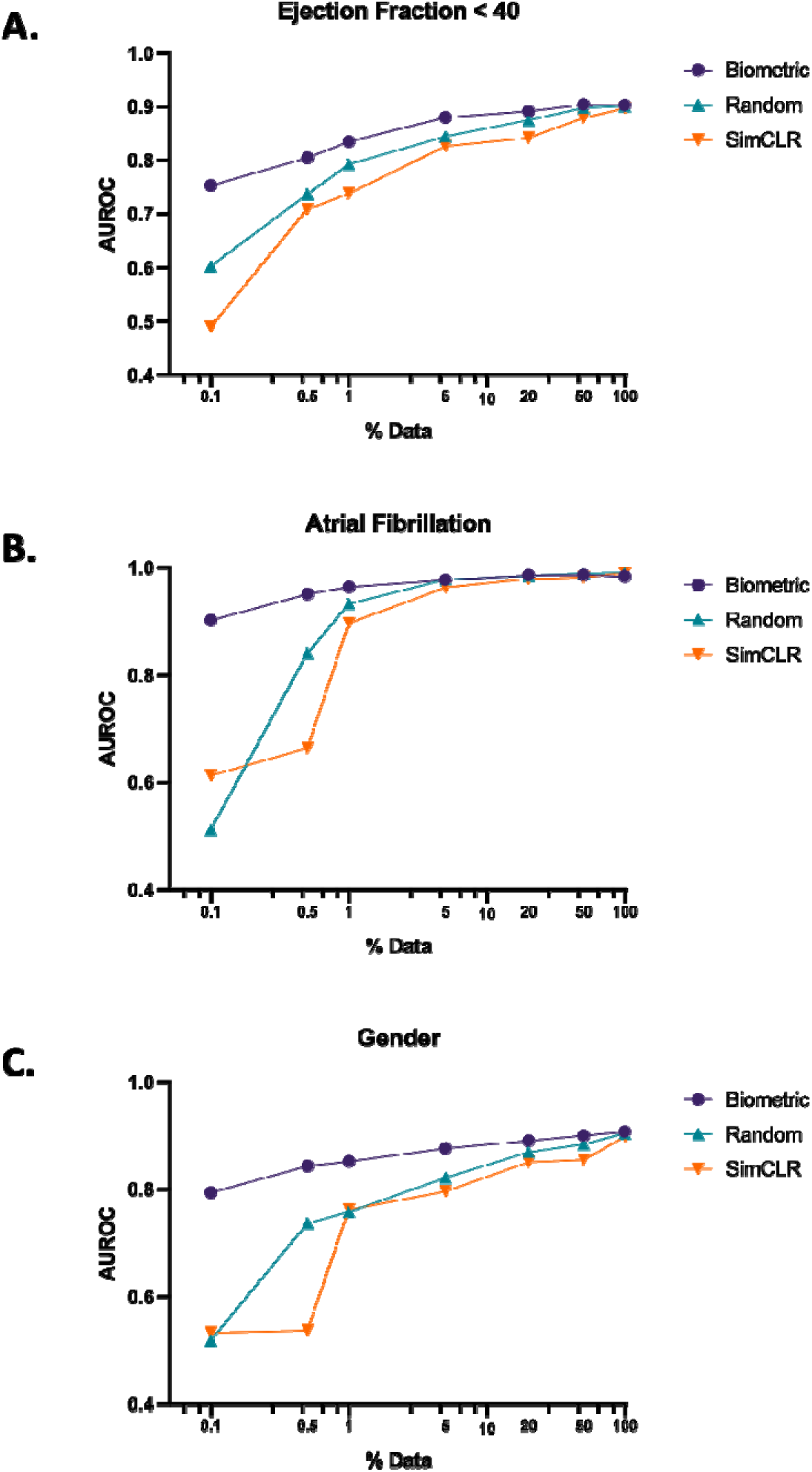
AUROC Curves in Held-Out Test Sets.

**Figure 3.**
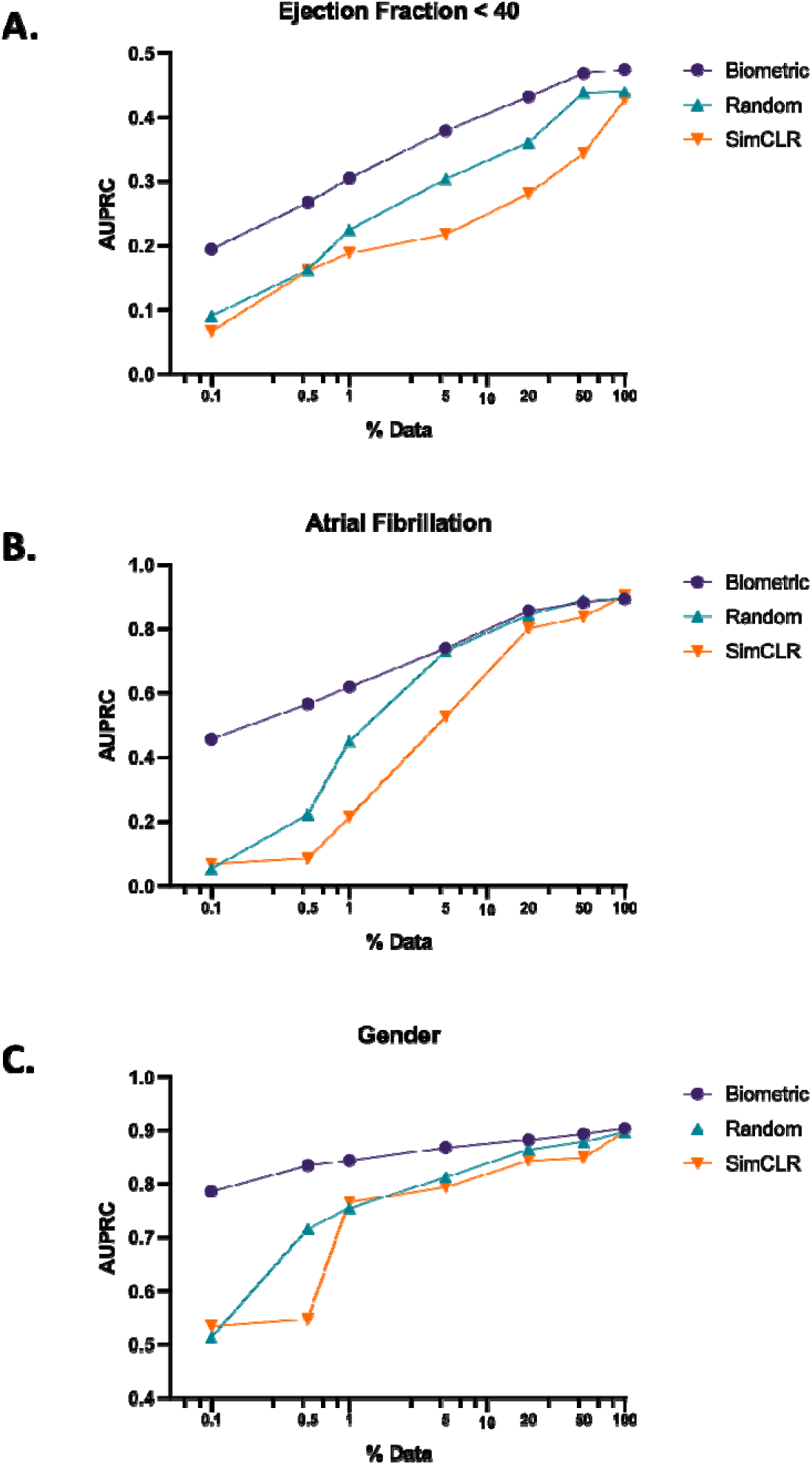
AUPRC Curves in Held-Out Test Sets.

As the quantity of data available progressively decreased, models trained using all three strategies saw lower performance with smaller size of the labeled training data, but BCL consistently outperformed other methods, with the difference in performance between BCL and other methods growing as labeled data became scarcer (**Figures 2 and 3**). On models trained with 1% of the available data, AUROC for models trained with BCL on the tasks of detecting LVSD, AF, and gender was 0.84, 0.96, and 0.85, respectively, while AUROC for randomly initialized models was 0.79, 0.93, and 0.76 respectively. AUROC for models trained with SimCLR was 0.74, 0.90, and 0.76 respectively. This corresponded to a mean gain in AUROC of 0.07 (8.7%) between BCL and the other two methods across tested applications (**Figure 4a**).

**Figure 4.**
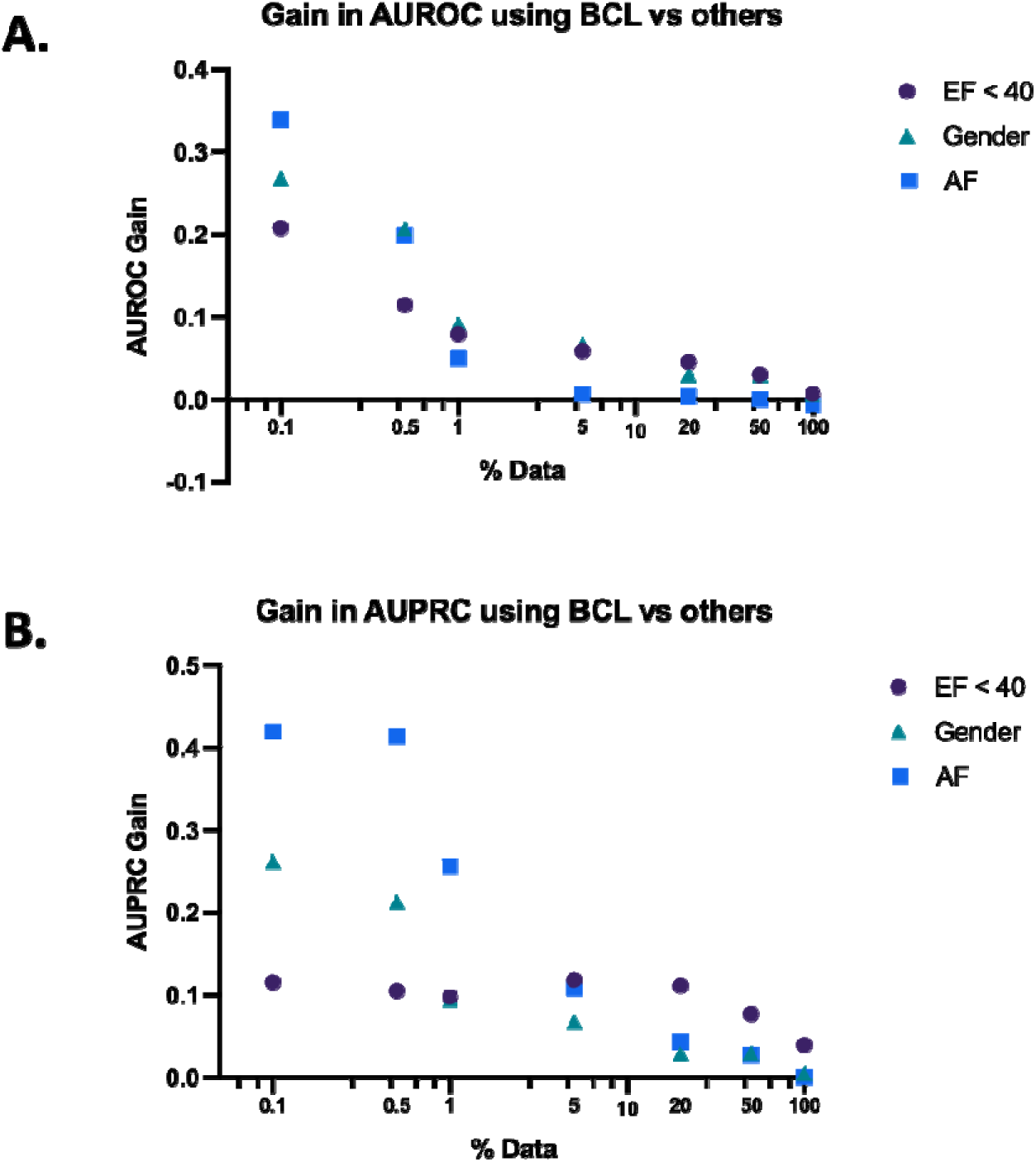
AUROC and AUPRC Gains in Held-Out Test Set.

On models trained with 1% of the data available, AUPRC for models trained with BCL on the tasks of detecting LVSD, AF, and gender was 0.31, 0.66, and 0.85 respectively, while AUPRC for randomly initialized models was 0.23, 0.55, and 0.75 respectively, and AUPRC for models trained with SimCLR was 0.19, 0.26, and 0.76 respectively. This corresponded to a mean gain in AUPRC of 0.15 (32.8%) between BCL and the other two methods across tested applications (**Figure 4b**).

On models trained with the smallest fraction of data available, 0.1%, AUROC for models trained with BCL on the tasks of detecting LVSD, AF, and Gender was 0.75, 0.90, and 0.79 respectively, and AUPRC was 0.19, 0.48, and 0.79. AUROC for randomly initialized models was 0.60, 0.51, and 0.52 respectively, and AUPRC was 0.09, 0.05, and 0.52. Finally, AUROC for models trained with SimCLR was 0.49, 0.61, and 0.53 respectively, and AUPRC was 0.07, 0.07, and 0.53. This corresponded to a mean gain in AUROC of 0.27 (49.7%) and AUPRC of 0.27 (119.8%) between BCL and the other two methods across tested applications (**Figure 4**).

### Performance in External Validation Datasets

The patterns observed in the held-out test set at small fractions of training dataset available were also observed in the validation datasets. On models trained on 1% of the data available, AUROC for models trained with BCL on the tasks of detecting LVSD, AF, and gender in the two external validation sets was 0.81, 0.97, and 0.80 respectively. AUROC for randomly initialized models was 0.70, 0.87, and 0.71 respectively, and AUROC for models trained with SimCLR was 0.66, 0.82, and 0.71 respectively (**Figure 5**). This corresponded to a mean gain in AUROC of 0.11 (15.3%) and AUPRC of 0.23 (42.8%) across applications between BCL and the other methods (**Figure 6**). There were similar patterns in AUROC and AUPRC when using other fractions of training data less than the entire dataset.

**Figure 5.**
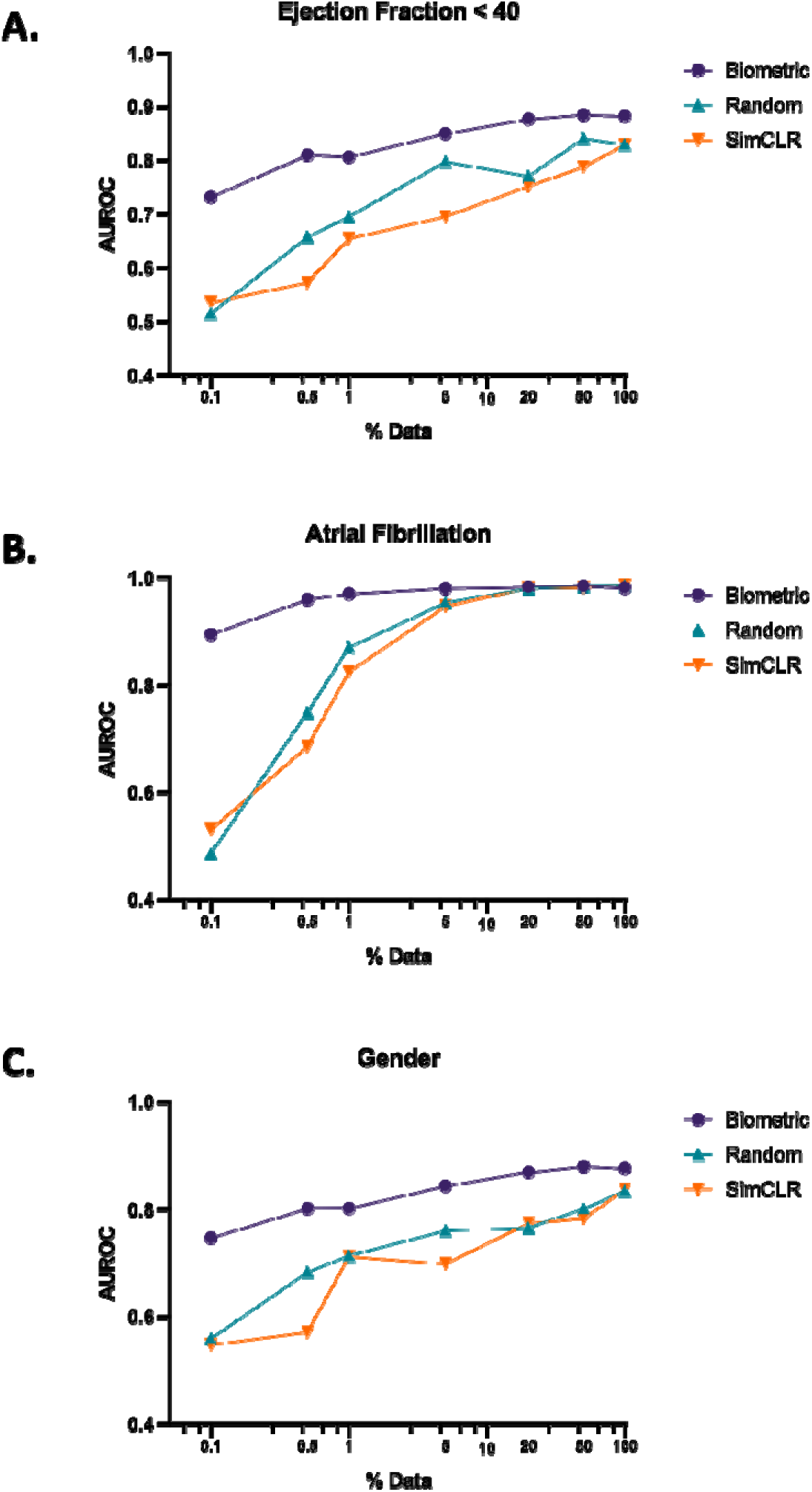
AUROC Curves in External Validation Sets.

**Figure 6.**
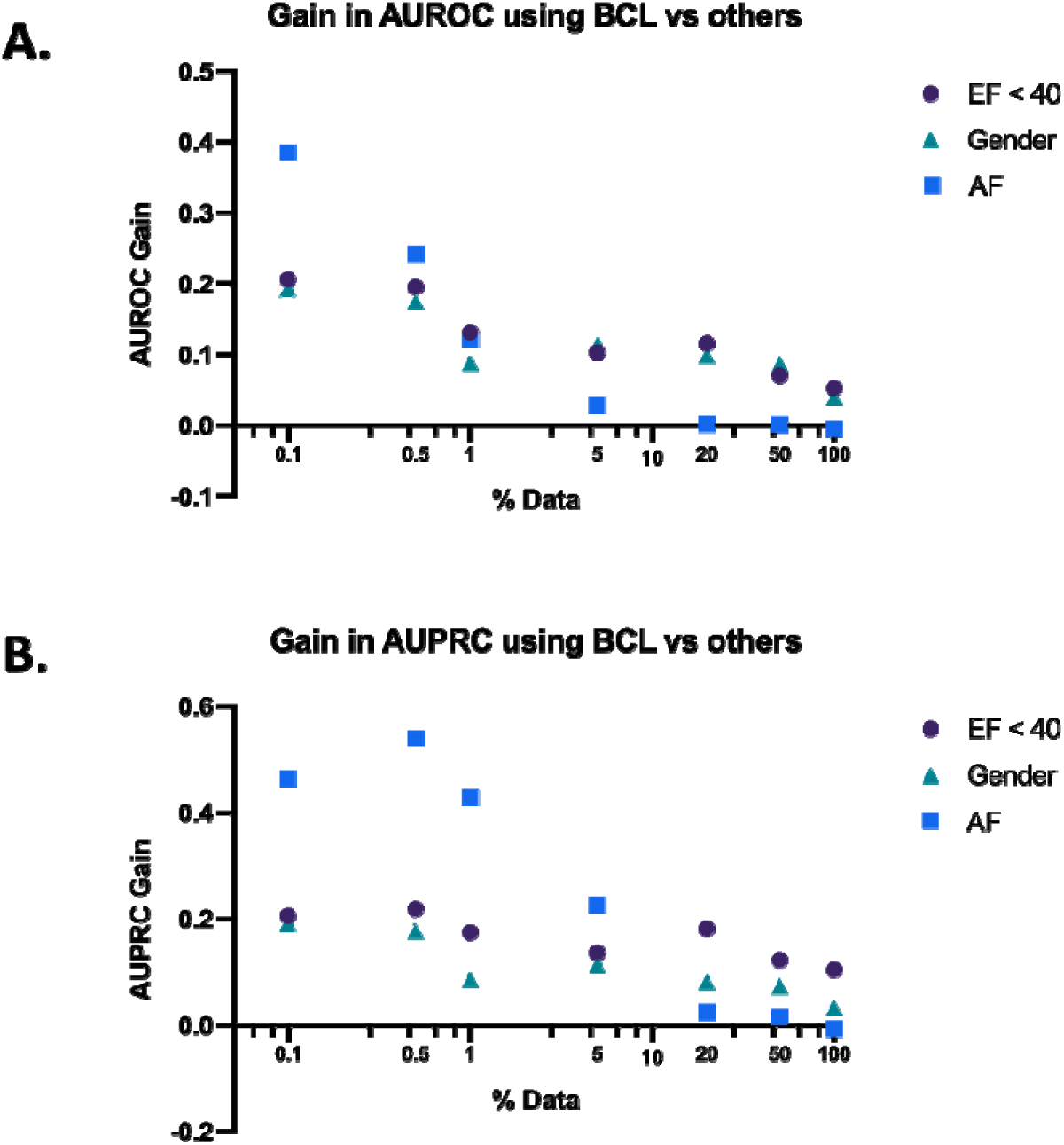
AUROC and AUPRC Gains in External Validation Sets.

On models trained with 100% of data available, BCL performed better in the external validation sets for the two hidden label tasks of LVSD and gender. AUROCs for models trained with BCL were 0.88 and 0.88 for LVSD and gender respectively, with random initializations were 0.83 and 0.83 respectively, and with SimCLR were 0.83 and 0.84 respectively (**Figure 6**). AUPRCs for models trained with BCL were 0.88 and 0.89 respectively, with random initializations were 0.75 and 0.85 respectively, and with SimCLR were 0.80 and 0.86 respectively (**Figure S2**). The initialization techniques demonstrated comparable performances for AF classification.

## DISCUSSION

Our novel biometric pretraining framework enables label-efficient learning on ECG images. The method focuses on building models to identify shared features of ECGs drawn from the same person at different times and plotted in different layouts and augmentations. This allows us to use unlabeled data to achieve large gains in the model development process for applications with sparsely labeled datasets, such as rare clinical disorders. While the model is not explicitly trained for any clinical diagnosis identification task during the pretraining process, it learns deeper representations from ECGs across different layouts. Using this method, a format-independent model for ECG images can be trained with just a few positive and negative examples.

Biometric Contrastive Learning (BCL) consistently outperforms two other commonly used initialization methods – random initialization and pretraining using SimCLR, the standard contrastive pretraining approach for image-based models. We compared the approaches on three tasks with differing biological bases – AF, gender, and LVSD. Evaluation across these 3 distinct tasks spanning clinical and hidden disorders demonstrated the broad relevance of this pretraining strategy for many discrete classification tasks. The fact that BCL generalized as the best method across all tasks indicates that it can be used across disease domains in the development of ECG models. BCL outperformed other methods by a larger margin as the quantity of data available decreased. Additionally, BCL performed better for hidden label tasks in our external validation datasets even when trained on 100% of the data. This suggests that the method is effective for both rare disorders when only a few positive examples are available for model development and may learn patterns that generalize better to new data sources even when trained with larger databases.

For random initializations, performances dropped across the three tasks as the amount of available training data became smaller, with drops in performance becoming larger as the scarcity of data increased. While BCL minimized this drop, SimCLR did not provide a performance boost for the three tasks studied, as has previously been reported in other medical image classification tasks^9,17^. SimCLR has effectively been applied for medical image tasks that are done by humans and are resilient to transformations like rotations, flips, and contortions of the image. ECG classification by humans relies on patterns in the plotted signals that are not as resilient to image contortions and may require a learned representation of patterns across several leads and time points in the image. It is possible that the representations of ECG images learned through the distortions in SimCLR are not relevant to the features of the image that contain information about both clinical labels diagnosable by physicians as well as hidden labels. On the other hand, it appears that the signatures stored in the BCL model, which learns which features of the image are unique to any given individual are indeed relevant to features useful for such classification tasks.

Our study has certain limitations that merit consideration. First, while the results suggest that BCL is most effective when training models for rare disorders, we do not compare pretraining strategies for a rare disorder, instead choosing to train models on fractions of labeled datasets with more common labels. This was essential to identify the threshold for training data. While our work indicates that BCL would be effective when used on rarer disorders, further investigation on other such disorders is necessary. Second, our models were built on a single CNN architecture, without an explicit evaluation against alternative models and architectures, and whether the pretraining strategy is more suitable for some models vs others. While the methods presented here should be generalizable to any encoder, future studies could evaluate these other architectures. Finally, our models were developed in a single institution. However, we have a large, diverse populations, and the models explicitly validated at other institutions and data sources. The ECG datasets are similar across sites, and therefore, our findings would suggest that the pretraining procedures would generalize to other sites developing models for ECG images.

## CONCLUSION

We developed a novel pretraining strategy that leverages the biometric signature of different ECGs from the same patient to significantly enhance data efficiency in developing AI-ECG models for ECG images, across several discrete tasks. This approach broadens the applications of image AI-ECG to rare disorders, for which training data is often limited, representing a significant advance in format-independent deep learning for the detection of heart disease directly from ECG images.

## FUNDING SOURCES

This study was supported by research funding awarded to Dr. Khera by the Yale School of Medicine and grant support from the National Heart, Lung, and Blood Institute of the National Institutes of Health under the award K23HL153775. The funders had no role in the design and conduct of the study; collection, management, analysis, and interpretation of the data; preparation, review, or approval of the manuscript; and decision to submit the manuscript for publication.

## CONFLICT OF INTEREST DISCLOSURES

Dr. Mortazavi reported receiving grants from the National Institute of Biomedical Imaging and Bioengineering, National Heart, Lung, and Blood Institute, US Food and Drug Administration, and the US Department of Defense Advanced Research Projects Agency outside the submitted work; in addition, Dr. Mortazavi has a pending patent on predictive models using electronic health records (US20180315507A1). Mr. Sangha and Dr. Khera are the coinventors of U.S. Provisional Patent Application No. 63/346,610, “Articles and methods for format-independent detection of hidden cardiovascular disease from printed electrocardiographic images using deep learning”. Dr. Khera receives support from the National Heart, Lung, and Blood Institute of the National Institutes of Health (under award K23HL153775) and the Doris Duke Charitable Foundation (under award 2022060). He receives support from the Blavatnik Foundation through the Blavatnik fund for Innovation at Yale. He also receives research support, through Yale, from Bristol-Myers Squibb, and Novo Nordisk. He is an Associate Editor at JAMA. In addition to 63/346,610, Dr. Khera is a coinventor of U.S. Provisional Patent Applications 63/177,117, 63/428,569, and 63/484,426. He is also a founder of Evidence2Health, a precision health platform to improve evidence-based cardiovascular care.

## Data Availability

The data cannot be shared publicly.

## ACKNOWLEDGEMENTS

**Author contributions:** RK conceived the study and accessed the data. VS developed the models. VS pursued the statistical analysis. VS drafted the manuscript. All authors provided feedback regarding the study design and made critical contributions to writing of the manuscript. RK supervised the study, procured funding, and is the guarantor.

## Notes

### Author Declarations

The study was reviewed by the Yale Institutional Review Board, which approved the study protocol and waived the need for informed consent as the study represents secondary analysis of existing data.

