## Supplemental File for "Biometric Contrastive Learning for Data-Efficient Deep Learning from Electrocardiographic Images"

### **SUPPLEMENTAL MATERIAL**

**(Sangha et al.)**

**Table S1. Analytic packages and language used for model development and statistical analysis**

| **Programming Language/Package** | **Version** |
| --- | --- |
| Python | 3.9.5 |
| TensorFlow | 2.8.0 |
| scikit-learn | 0.24.2 |
| pandas | 1.3.1 |
| numpy | 1.19.5 |

#### **Figure S1. EfficientNet-B3 architecture used for model development.** Our image-based convolutional neural network (CNN) is designed Efficientnet-B3 architecture to recognize visual patterns of LV systolic dysfunction from ECG-images. The input layer receives the ECG image as a matrix of pixel values. The convolutional layer applies a set of learnable filters to the input image that slide across the image, convolving it and producing an output feature map. These filters detect different features, such as edges, shapes, or textures, that are important for identifying patterns in the ECG image. The pooling layer reduces the spatial size of the output feature maps by performing a down-sampling operation, which helps to make the model more robust to variations in the image. Finally, the output layer produces the prediction of LV systolic dysfunction from the image.

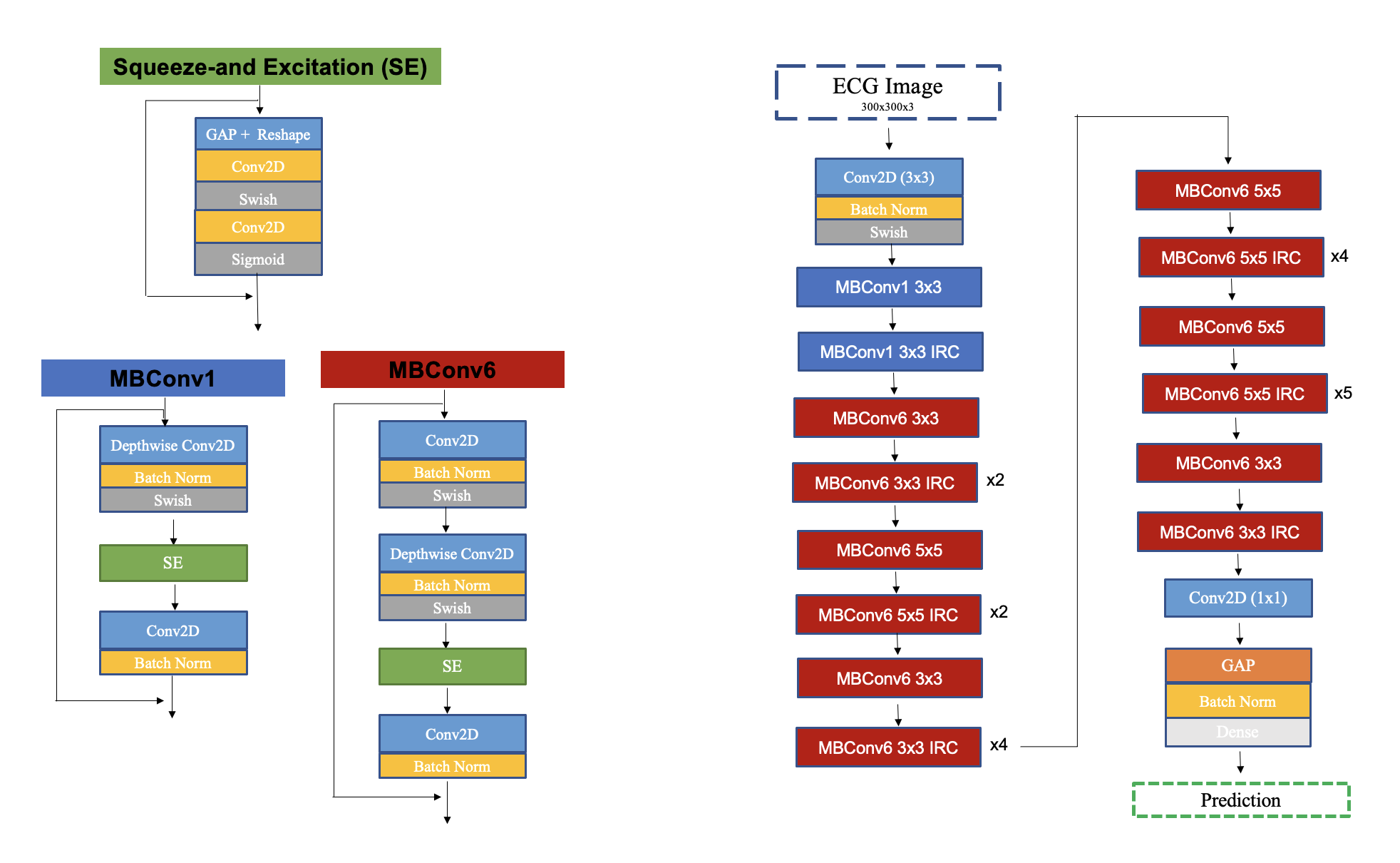

**Figure S2. AUPRC Curves in External Validation Sets**

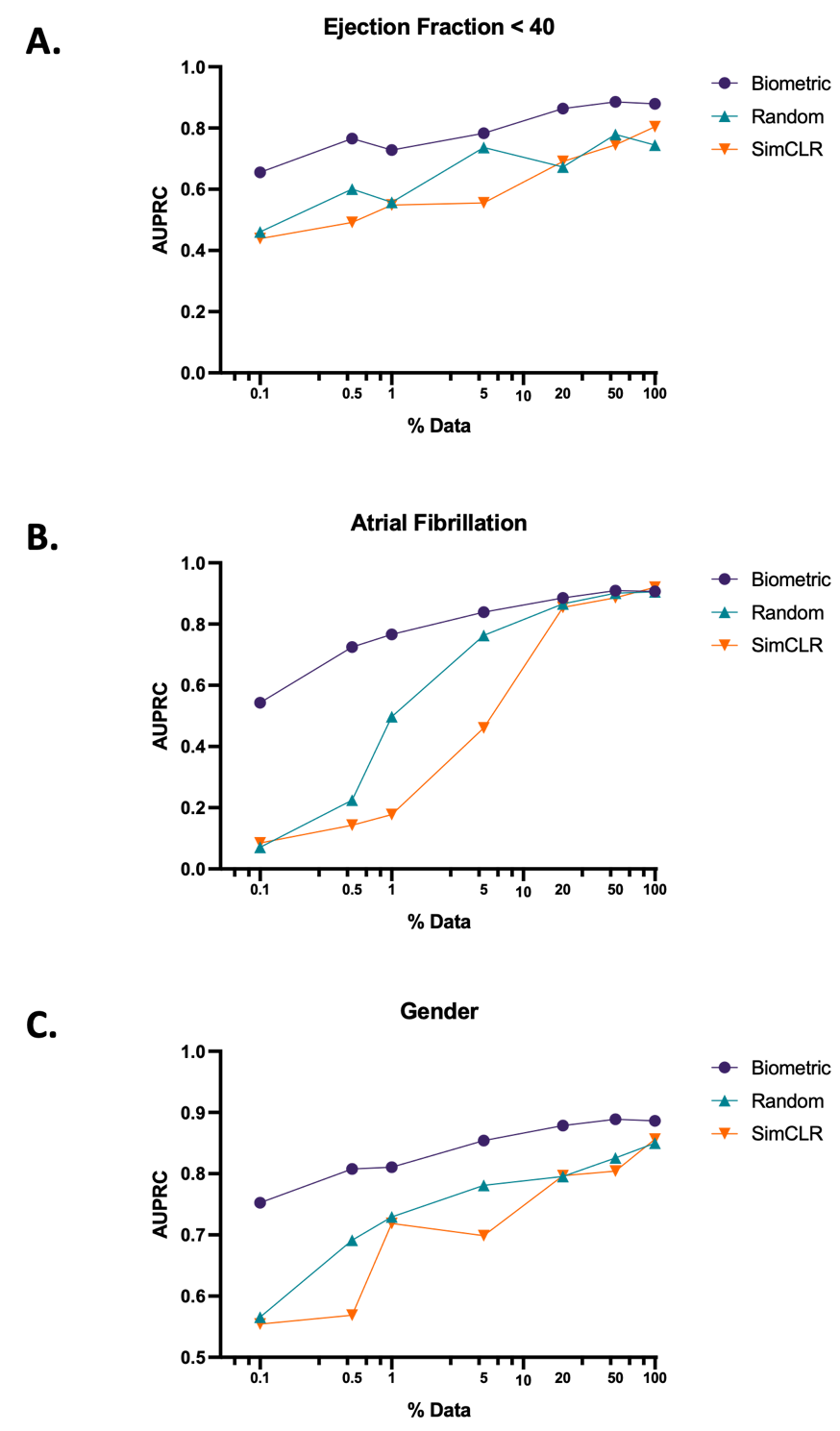
